# Short-term effects of BNT162b2 mRNA COVID-19 vaccination on physiological measures: a prospective study

**DOI:** 10.1101/2021.05.06.21256587

**Authors:** Yftach Gepner, Merav Mofaz, Shay Oved, Matan Yechezkel, Keren Constantini, Nir Goldstein, Erez Shmueli, Dan Yamin

**Affiliations:** Department of Epidemiology and Preventive Medicine, School of Public Health, Sackler Faculty of Medicine, and Sylvan Adams Sports Institute, Tel-Aviv University, Tel-Aviv, Israel; Department of Industrial Engineering, Tel-Aviv University, Tel-Aviv, Israel; Center for Combatting Pandemics, Tel-Aviv University, Tel-Aviv, Israel

## Abstract

**Background:** Clinical trial guidelines for assessing the safety of vaccines, including the FDA criteria, are primarily based on subjective, self-reported questionnaires. Despite the tremendous technological advances in recent years, objective, continuous assessment of physiological measures post-vaccination is rarely performed.

**Methods:** To evaluate the short-term effects of the BNT162b2 COVID-19 vaccine on physiological measures, we conducted a prospective observational study during the mass vaccination campaign in Israel. 160 individuals >18 years who were not previously found to be COVID-19 positive and who received the second dose of the COVID-19 vaccine between 1 January, 2021, and 13 March, 2021 were equipped with a chest-patch sensor and a dedicated mobile application. The chest-patch sensor continuously measured 13 physiological vitals one day before the inoculation (baseline), for four days: heart rate, blood oxygen saturation, respiratory rate, systolic and diastolic blood pressure, pulse pressure, mean arterial pressure, heart rate variability, stroke volume, cardiac output, cardiac index, systemic vascular resistance, and body temperature. The mobile application collected daily self-reported questionnaires starting one day before the inoculation, for 15 days on local and systemic reactions, sleep quality, stress levels, physical activity, and mood levels.

**Findings:** Within the first 48 hours post-vaccination, we identified significant changes (p-value <0.05) in nearly all 13 chest-patch indicators compared to their baseline levels. 48.5% (n=78) reported no local or systemic reaction. Nevertheless, we identified considerable changes in chest-patch indicators during the first 48 hours post-vaccination also in this group of presumably asymptomatic participants. Within three days from vaccination, these measures returned to baseline levels in both groups, further supporting the safety of the vaccine.

**Interpretation:** Our work underscores the importance of obtaining objective physiological data in addition to self-reported questionnaires when performing clinical trials, particularly in ones conducted in very short time frames.

**Funding:** The European Research Council (ERC) project #949850.

## Introduction

Vaccination is widely accepted as the most prominent measure in the fight against COVID-19, posing the greatest hope for ending this major global health pandemic and related economic crisis.^1,2^ Consequently, an unprecedented international effort by private and public institutions alike was directed at accelerating the traditionally lengthy vaccine-development process.^3–5^ On 2 December 2020, less than a year from the pandemic outbreak, the first vaccine, BNT162b2 mRNA (Pfizer-BioNTech), was granted an Emergency Use Authorization (EUA) by the UK Medicines and Healthcare products Regulatory Agency (MHRA).^6^ This initial authorization was followed by rapid authorizations for emergency use in several countries, with the US Food and Drug Administration (FDA) among the first to do so.^7^ The promising BNT162b2 vaccine was demonstrated to have 95% efficacy in preventing symptomatic COVID-19 in clinical trials,^8^ and 92% efficacy in a nationwide mass vaccination.^9^

Safety data from a randomized, controlled trial suggests a favorable safety profile for the BNT162b2 vaccine.^8^ Specifically, the local and systemic reactions reported during the first seven days after vaccination were mostly mild to moderate, with a median onset of 0–2 days after vaccine administration and a median duration of 1–2 days. The most frequently reported reactions were fatigue, headache, muscle pain, chills, joint pain, and fever. The incidence of serious adverse events was low and was similar between vaccine- and placebo-treated participants. The safety of the new vaccine over a median of two months post-vaccination was similar to that of other viral vaccines. A considerable fraction of the participants did not report any reaction or adverse event. Likewise, several other vaccine candidates, including ChAdOx1 nCoV-19 (Oxford/AstraZeneca) and mRNA-1273 (Moderna), received EUAs following similar encouraging safety results in randomized, controlled trials.^10–12^

Nevertheless, concerns regarding potential adverse effects from vaccines have recently led to the suspension of the ChAdOx1 nCoV-19 vaccination campaigns in several European countries,^13^ and may have reinforced the public hesitancy towards COVID-19 vaccines. These concerns underscore the importance of extracting as much information as possible from clinical trials. However, to date, clinical trial guidelines for assessing the safety of vaccines, including the FDA criteria,^14^ are primarily based on subjective, self-reported questionnaires. Despite the tremendous technological advances in recent years, objective, continuous assessment of physiological measures post-vaccination is rarely performed.

Here, we evaluated the short-term effects of the BNT162b2 COVID-19 vaccine on physiological measures. We followed a cohort of 160 participants who received the second dose of the BNT162b2 vaccine for 96 hours, from 24 hours prior to vaccine administration until 72 hours after the inoculation. Participants were fit with a chest-patch sensor that monitored objectively and continuously 13 different physiological indicators. Additionally, a dedicated mobile application was used to record daily self-reported questionnaires on local and systemic reactions and various well-being indicators. We identified considerable changes in chest-patch indicators during the first 48 hours post-vaccination also in this group of presumably asymptomatic participants. These measures returned to baseline levels in both groups, further supporting the safety of the vaccine. Our findings underscore the importance of accounting for objective technological advances in clinical trials to more accurately understand the invisible impact of the vaccine on our respiratory, cardiovascular, and hemodynamic systems. Extracting as much information as possible from clinical trials, particularly during an emergency, is crucial for a more comprehensive determination of vaccine safety.

## Materials and Methods

### Study design and participants

Our study includes a prospective cohort of 160 participants who were not previously found to be COVID-19 positive and received the second dose of the BNT162b2 mRNA COVID-19 vaccine between 1 January, 2021, and 13 March, 2021 (study protocol is available at the appendix pp 25-35). As reported reactions were considerably more severe after the second dose compared to the first,^8^ we focused on the second dose of vaccine. Out of the 160 participants in this study, 90 (56.25%) were women, and 70 (43.75%) were men. Their age ranged between 21 and 78 years, with a median age of 40. Participants were equipped with a chest-patch sensor and were monitored for a period of four days, starting one day prior to vaccine administration. In addition, participants installed a dedicated mobile application and were requested to fill in a daily questionnaire, starting one day prior to the inoculation, for a period of 15 days. For each participant, the 24-hour period prior to vaccination served as a baseline.

In order to recruit participants and ensure they complete all the study’s requirements, we hired a professional survey company. Potential participants were recruited through advertisements in social media, online banners, and word-of-mouth. The survey company was responsible for guaranteeing the participants met the study’s requirements, in particular, that they agreed to wear the chest-patch sensor and fill in the daily questionnaires.

Participants were met in person, roughly 24 hours prior to vaccination, and received a detailed explanation about the study, after which they were requested to sign an informed consent form. Then, participants were asked to complete a one-time enrollment questionnaire and install two applications on their mobile phones: an application that passively collects data from the chest-patch sensor and the PerMed application, allowing participants to fill the daily questionnaires.

To better understand the effects of the second vaccine dose on physiological measures, we also monitored a subset of 25 participants, with an identical procedure, when receiving their first vaccination dose.

### The chest-patch sensor

The photoplethysmography (PPG)-based chest monitors purchased and used in this study collects the following 13 indicators of vital signs: heart rate, blood oxygen saturation, respiratory rate, systolic and diastolic blood pressure, pulse pressure, mean arterial pressure, heart rate variability, stroke volume, cardiac output, cardiac index, systemic vascular resistance (SVR) and body temperature. Together, these indicators of vital signs provide a comprehensive assessment of the respiratory, cardiovascular, and hemodynamic state. The chest patch sensor received FDA clearance for measures of heart rate, respiratory rate, and systolic and diastolic blood pressure (clearance number K190792) and CE Mark approval for all 13 measures (CE 2797) (Biobeat Technologies Ltd). To the best of our knowledge, this is the only cleared wearable wireless medical-grade device to provide all these measurements. The sensor tracks vital signs derived from changes in the pulse contour, following baseline calibration using an approved non-invasive, cuff-based device, and is based on Pulse Wave Transit Time (PWTT) technology, combined with Pulse Wave Analysis (PWA) (see, for example, ^15,16^). The sensor continuously collected data at 10-minute intervals for the entire duration of the 96-hour experiment.

### PerMed mobile application

Developed originally to support the PerMed study,^17^ the PerMed mobile application passively collects smartphone sensory data, as well as allows participants to fill the daily questionnaires. The daily questionnaire we used included questions about: mood level (on a scale of -2 [awful] to 2 [excellent]), stress level (on a scale of -2 [very low] to 2 [very high]), sleep quality (on a scale of -2 [awful] to 2 [excellent]), physical activity (in minutes), clinical symptoms (from a closed list of local and systemic reactions observed in the BNT162b2 mRNA Covid-19 clinical trial,^8^ with an option to add other symptoms as free text).

In order to improve the quality and reliability of the data and to ensure its continuous collection, we applied the following two measures: 1) Participants who did not complete the daily questionnaire by 7 pm received a notification in their mobile app to fill the questionnaire; 2) We developed a dedicated dashboard that helped us identify when participants did not fill in the daily questionnaires. Those participants were contacted by the survey company and were encouraged to cooperate better.

### Statistical analysis

Before analyzing the data, we performed several preprocessing steps. With regard to the daily questionnaires, in cases where participants filled in the daily questionnaire more than once on a given day, only the last entry for that day was considered, as it was reasoned that the last one likely best represented the entire day. Self-reported symptoms that were entered as free text were manually categorized. With regard to the chest-patch indicators, data were first aggregated per hour (by taking the mean value). Then, to impute missing values, we performed a linear interpolation.

To examine the changes in each chest-patch indicator over the 72 hours post-vaccination compared with the 24-hour baseline period prior to vaccination, we performed the following steps. For each indicator, for each hour *h* of the 72 hours post-vaccination, we calculated for each individual the mean value in the five-hour sliding window: [*h*−4, *h*−3, *h*−2, *h*−1, *h*]. Then, we calculated the relative change in the percentage of this value compared to the corresponding five-hour window in the baseline period. Finally, we calculated the mean value for hour *h* over all 160 participants, as well as the 90% confidence interval, corresponding to a significance level of 0.05 in a one-sided t-test (figure 1). We performed a similar analysis for the 25 participants who received the first vaccine dose (appendix pp 44-45).

**Figure 1.**
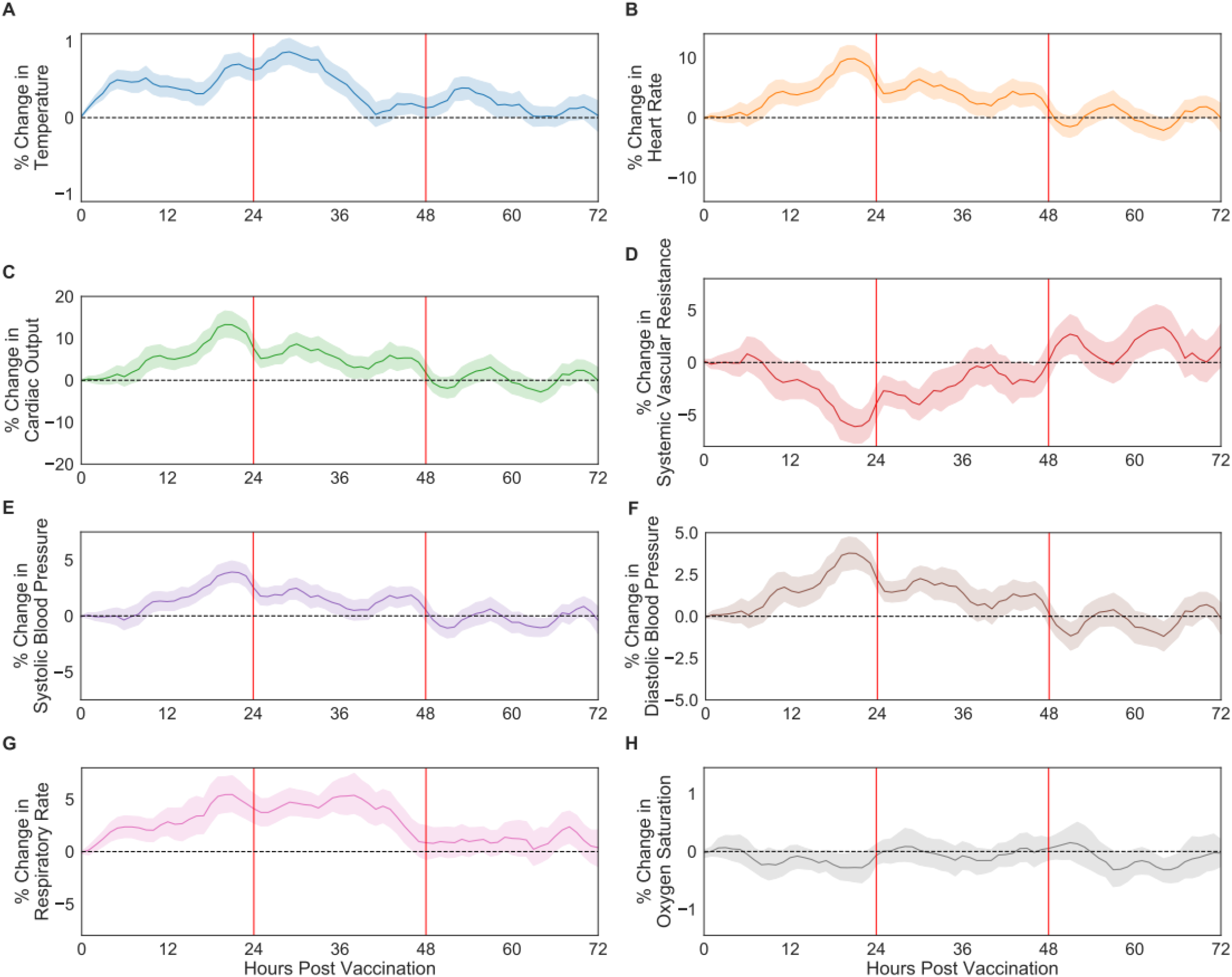
Percentage of change in respiratory, cardiovascular, and physiological indicators recorded by the chest-patch sensor compared to their baseline levels: (A) body temperature, (B) heart rate, (C) cardiac output, (D) systemic vascular resistance, (E) systolic blood pressure, (F) diastolic blood pressure, (G) respiratory rate, and (H) oxygen saturation. Mean values are depicted as solid lines, 90% confidence intervals are presented as shaded regions, and horizontal dashed lines represent no change compared to the baseline levels.

Next, for each of the three days after the vaccination, we calculated the percentage of participants who reported new local or systemic reactions compared to their baseline period. For each reaction, a 90% confidence interval was calculated assuming a beta distribution, with parameter *α* corresponding to the number of participants reporting that reaction plus one (i.e., “successes”), and parameter *β* corresponding to the number of participants who did not report that reaction plus one (i.e., “failures”).

Similarly, we calculated the changes in the well-being indicators reported post-vaccination compared to those reported in the baseline period. Specifically, for each indicator, we calculated the difference between the value in each day and the corresponding value in the baseline period. Then, we calculated the mean difference value over all participants and a 90% confidence interval using a *t* distribution.

Finally, we examined the difference between symptomatic and asymptomatic participants with regard to changes in the chest-patch indicators, stratified by the number of days post-vaccination (1-3) and part of the day (day or night) (Figure 3). For a given day post-vaccination, symptomatic participants were defined as those who reported at least one reaction on that day that they did not report in the baseline period. Asymptomatic individuals were defined as those who reported no reactions on that day. We defined nighttime as the time interval between 12:00 am and 7:00 am and daytime between 7:00 am and 12:00 am. This day-night definition is consistent with the observed movement patterns of the participants throughout the study. For each participant, we calculated the mean indicator value for each day and part of the day post-vaccination. Then, we calculated the relative change in percentages of these values compared to their corresponding values in the baseline period. Next, we calculated the mean values of symptomatic participants and asymptomatic participants, as well as their corresponding 90% confidence intervals using a *t* distribution. Finally, unequal variances *t*-tests were used to evaluate the differences between symptomatic and asymptomatic participants.

**Figure 2.**
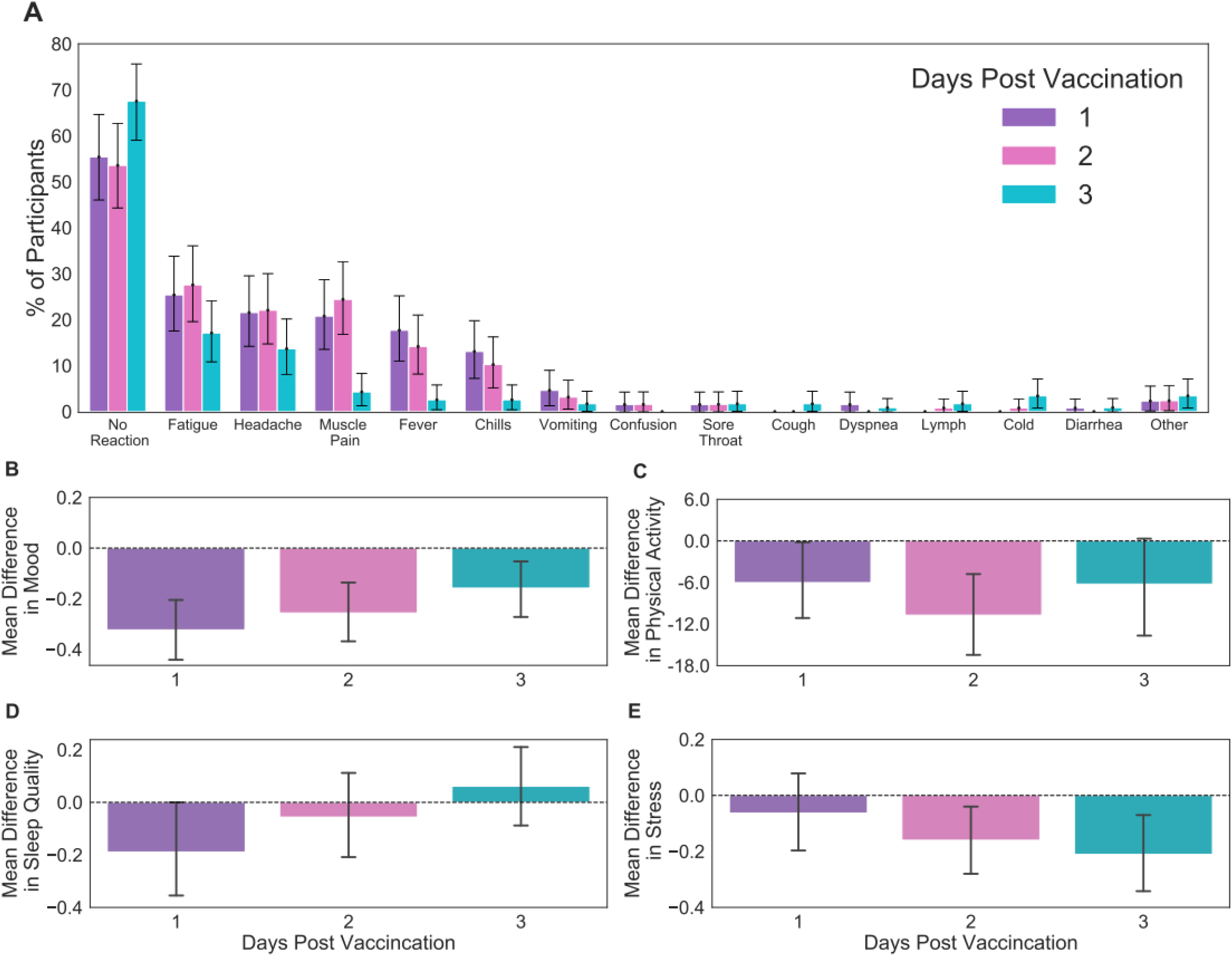
Local and systemic reactions and well-being indicators reported by participants through the mobile application. (A) local and systemic reactions, (B) mood level, measured on a 1 to 5 Likert scale, (C) duration of physical activity, measured in minutes, (D) sleep quality, measured on a 1 to 5 Likert scale, and (E) stress level, measured on a 1 to 5 Likert scale. Well-being indicators were calculated by subtracting the baseline values from the daily values. Error bars represent 90% confidence intervals. Horizontal dashed lines represent no change compared to the baseline levels.

**Figure 3.**
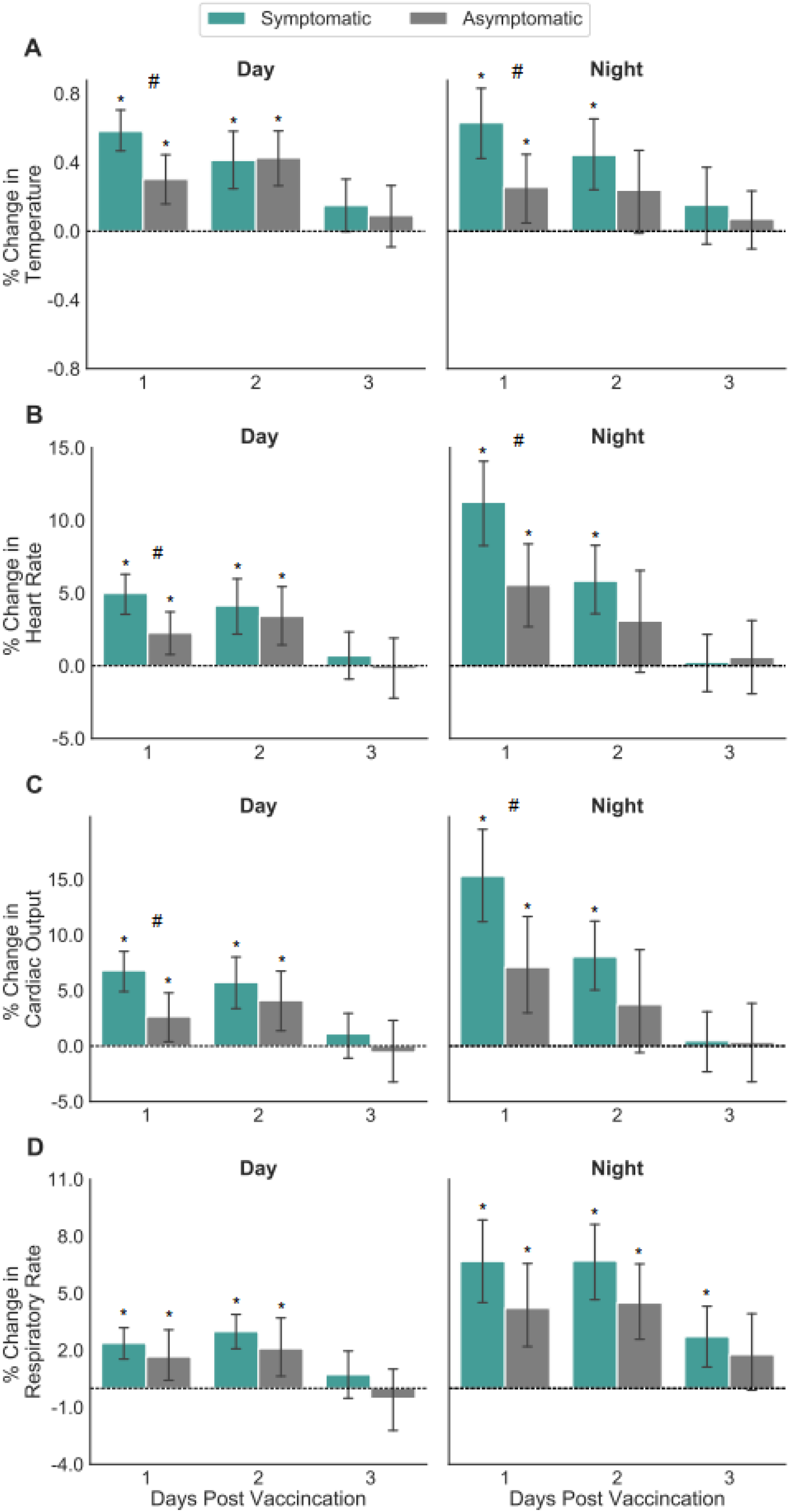
Percentage of change in chest-patch indicators for participants who reported at least one local or systemic reaction (symptomatic) and those who reported no reaction (asymptomatic) during the daytime and the nighttime: (A) body temperature, (B) heart rate, (C) cardiac output, and (D) respiratory rate. Error bars represent 90% confidence intervals. Horizontal dashed lines represent no change compared to the baseline levels. Significant differences from baseline at a 0.05 level are marked with *. Significant differences between symptomatic and asymptomatic participants at a 0.05 level are marked with #.

### Ethical Approval

Before participating in the study, all subjects were advised, both orally and in writing, as to the nature of the study and gave written informed consent to the study protocol (appendix pp 25-35), which was approved by the Tel-Aviv University Institutional Review Board (0002522-1). All data were de-identified, and no personal identifiable information was gathered.

## Results

Between 1 January 2021, and 13 March 2021, a total of 166 participants were recruited. Among them, 160 participants completed the trial, and their data were analyzed; five participants left the trial early, and one participant was provided with a malfunctioning chest-patch sensor. Among these participants, 56.25% were females, 13.75% were obese (body-mass index of at least 30.0), 18.12% had high blood pressure, and 20.00% had at least one comorbidity. The median age was 40 years, and 10.63% of participants were older than 59 years of age (Table 1).

**Table 1.** Characteristics of the participants

| **Characteristic** | **Number of participants (%)**<br>**(N=160)** |
| --- | --- |
| **Gender** |  |
| Male | 70 (43.75%) |
| Female | 90 (56.25%) |
| **Age group** |  |
| 18-39 yr | 91 (56.87%) |
| 40-59 yr | 52 (32.50%) |
| ≥ 60 yr | 17 (10.63%) |
| **Body-mass index\*** |  |
| < 30.0 | 128 (80.00%) |
| ≥ 30.0 | 22 (13.75%) |
| Unspecified | 10 (6.25%) |
| **High blood pressure measured at baseline (Systolic >140 or Diastolic >90)\*\*** |  |
| No | 131 (81.88%) |
| Yes | 29 (18.12%) |
| **Household size** |  |
| ≤ 2 | 67 (41.87%) |
| > 2 | 83 (51.88%) |
| Unspecified | 10 (6.25%) |
| **Self-reported illness in the past 30 days** |  |
| No | 104 (65.00%) |
| Yes | 46 (28.75%) |
| Unspecified | 10 (6.25%) |
| **Comorbidities** |  |
| No | 128 (80.00%) |
| Hypertension | 11 (6.87%) |
| Diabetes | 7 (4.37%) |
| Heart disease | 3 (1.87%) |
| Chronic lung disease | 5 (3.12%) |
| Renal failure | 1 (0.62%) |
| Unspecified | 12 (7.50%) |

\* The body-mass index is the weight in kilograms divided by the square of the height in meters
\*\* Blood pressure was measured a day before inoculation

Within the first 48 hours post-vaccination, we identified significant changes in nearly all 13 chest-patch indicators compared to their baseline levels (figure 1 and appendix p 36). For example, at their peak, the heart rate increased by 9.85% (90% CI, 7.71% to 11.99%), the systolic blood pressure increased by 3.91% (90% CI, 2.97% to 4.87%), and the diastolic blood pressure increased by 3.78% (90% CI, 2.82% to 4.74%), compared to baseline levels. By contrast, we observed no significant differences in blood oxygen saturation (figure 1B). Following the initial 48 hours, these changes faded, with measurements returning to their baseline levels.

Focusing on self-reported reactions, we find consistent trends with those observed in the BNT162b2 mRNA vaccine clinical trial (figure 2A and appendix pp 37-38). Specifically, the most frequent reactions reported via the self-reported questionnaires collected from the mobile application were fatigue, headache, muscle pain, fever, and chills. Consistent with our analysis of the chest-patch indicators, participants reported most of the reactions during the first two days post-vaccination, followed by a sharp decline in reporting on the third day. Importantly, almost half of the participants (48.5%) did not note any local or systemic reaction.

We also observe considerable changes in self-reported well-being indicators post-vaccination (figure 2B-E). During the first two days post-vaccination, participants exhibited a significant reduction in mood level (figure 2B), sleep quality (figure 2C), and time spent on physical activity (figure 2D) compared to baseline levels. Interestingly, participants reported significantly lower stress levels on the second- and third-day post-vaccination compared to the day prior to vaccination (figure 2E).

Importantly, we identified significant changes in chest-patch indicators during the first two days post-vaccination also in presumably asymptomatic participants (figure 3 and appendix pp 39-40). Specifically, during the daytime of the first two days post-vaccination, nearly all 13 chest-patch indicators significantly changed not only for symptomatic participants (i.e., those who reported at least one local or systemic reaction) but also for asymptomatic participants (i.e., those who did not report any local or systemic reaction). During the night hours, among the asymptomatic participants, significant changes were observed mainly on the first night post-vaccination. Conversely, among the symptomatic participants, significant changes were observed on both the first- and second-nights post vaccination. Moreover, the changes observed for symptomatic participants were found to be significantly higher than those of asymptomatic individuals in 9 out of the 13 chest-patch indicators during the first day post-vaccination and in 8 out of the 13 chest-patch indicators during the first night post-vaccination (unequal variances *t*-test, *p*-value<0.05).

We also analyzed the changes in chest-patch indicators compared to baseline levels, stratified by age group, gender, and vaccine dose. Participants 60 years old and above exhibited milder, albeit not significant, changes than those below 60 years old in nearly all chest-patch indicators (appendix pp 41-45). We found no differences between men and women in chest-patch indicators. In contrast to the second vaccine dose, examining the subset of 25 participants who were monitored when receiving their first vaccine dose revealed no difference in vitals during the first 48 hours post-inoculation (appendix pp 44-45).

## Discussion

Our key findings suggest that multiple physiological measures significantly increase following BNT162b2 vaccine administration in both participants who reported and those who did not report local and systemic reactions. Within three days from vaccination, these measures returned to baseline levels in both groups, further supporting the safety of the vaccine.

We identified several aspects that strengthen the short-term safety of the BNT162b2 vaccine. First, we found that all the physiological measures returned to their baseline levels within three days from vaccination. Second, we observed no change in oxygen saturation levels compared to their baseline levels, indicating that major adverse health consequences are less likely. Third, reports of local and systemic reactions declined considerably on the third day following vaccination. Fourth, well-being indicators reported by the participants, including mood, sleep quality, and physical activity, also returned to their baseline levels by the third day. Interestingly, the reported stress levels significantly improved on the second and third days after vaccination, suggesting that participants were concerned prior to vaccine administration. Thus, our study quells, in part, concerns raised by those hesitant to be vaccinated on the grounds of potential adverse vaccination consequences by demonstrating that such consequences are likely to fade after a few days.

We identified a considerable decline in SVR during the first two days after inoculation. Such a decline is typically regarded as an indication of inflammatory response. Hematology studies performed by Pfizer and reported in the VRBPAC document,^18^ suggest that the most commonly observed changes were transient grade 1 or 2 decreases in lymphocytes, one to three days after the first vaccine dose. These decreases returned to baseline levels within 6-8 days of the first dose. The authors note that RNA vaccines are known to induce type I interferon, which regulates lymphocyte recirculation and are associated with transient migration and redistribution of lymphocytes. This rapid rebound of lymphocytes supports the notion that they were not depleted, but temporarily migrated out of the peripheral blood and subsequently re-entered the bloodstream by the next assessment time. Although these hematology observations were after the first dose, the change in vascular resistance could provide compelling support for the idea of migration of lymphocytes out of circulation. However, our study did not include hematology tests. Though SVR is regarded as an indication of the inflammatory response, the fact it is counterbalanced with cardiac output, and the lack of other inflammatory indicators, including body temperature, show that the decrease is not necessarily directly related in this situation to inflammation. This could be further looked at in future studies, either with this specific vaccine or with any other, emphasizing the need to exhaust all available data in clinical research to detect – as early as possible – any side effects among all volunteers.

Our study includes several limitations. First, our cohort includes only 160 participants and may not adequately represent the vaccinated population in Israel and elsewhere. Nevertheless, despite this considerably small sample size, trends observed by the chest-patch sensor were found to be significant. Moreover, the proportion of those who reported local and systemic reactions and the type of reactions noted were similar to those observed in clinical trials ^8^. Second, all participants received the BNT162b2 vaccine, which was the only vaccine used in Israel. Given the similarities regarding the safety observed between different COVID-19 vaccines,^8,11,12^ we believe our findings are likely to be qualitatively similar in other vaccine types. Third, the rich data gathered by the chest-patch sensor was recorded for a relatively short period, four days. However, our findings from both the chest-patch sensor and the daily questionnaire (which was collected for a more extended period, i.e., 14 days) reveal that the vast majority of local and systemic reactions faded within two days. Fourth, we did not explicitly control for the effects of the clinical trial setting (i.e., participating in a trial, wearing a chest-patch sensor, potential concerns from the vaccine, etc.). While one may argue that the observed changes may be an artifact of the trial setting, we would expect to find these changes during the first vaccine dose as well. However, since we found no differences in most vitals than baseline levels in the subset of participants who received their first dose, we believe the changes observed after the second dose arises from an actual reaction to the vaccine.

About half of the participants (48.5%) did not report any local or systemic reaction following vaccination, which is consistent with previous reports from the clinical trial ^8^. Yet, we show that both symptomatic and asymptomatic participants had substantial objective physiological changes regardless of their subjective reports. Hence, our work underscores the importance of obtaining objective physiological data in addition to self-reported questionnaires when performing clinical trials, particularly in those conducted in very short time frames.

Whereas self-reported trends are widely described in the scientific literature, no study or vaccine clinical trial has reported the comprehensive effects of the COVID-19 vaccine on physiological measures. In fact, current US FDA, and European Medicines Agency (EMA) guidelines for assessing the safety of newly developed vaccines are primarily based on subjective, self-reported questionnaires. Our findings should encourage public health officials and regulatory agencies to include the analysis of objective measures, in addition to self-reported questionnaires, as part of the evaluation of clinical trials. Though not available in the past, current digital wearable health technologies could offer simple platforms for obtaining physiological measures, adding invaluable objective, non-biased information. Thus, as part of the future process of vaccine approval, it is essential to combine objective and remote medical-grade monitors in clinical studies.

## Research in context

### Evidence before this study

Clinical trial guidelines for assessing the safety of vaccines, including the FDA criteria, are primarily based on subjective, self-reported questionnaires. Despite the tremendous technological advances in recent years, objective, continuous assessment of physiological measures post-vaccination is rarely performed. We conducted an extensive literature review from 5-17 December 2020 and aimed to include all relevant research with no language restrictions. We searched PubMed, Scopus, and Google Scholars for eligible studies using keywords such as “COVID-19 vaccine”, “vaccine adverse effects”, “vaccine reactions”, “vaccine clinical trial”. We also checked the FDA guidelines and CE regulations regarding vaccine trails and approval.

### Added value of this study

To our knowledge, this study is the first to account for objective, continuous assessment of physiological measures post-vaccination. Our approach revealed considerable changes in 13 different measures following the BNT162b2 vaccine administration in both participants who reported and those who did not report local and systemic reactions. Moreover, within three days from vaccination, these measures returned to baseline levels in both groups, further supporting the vaccine’s safety.

### Implications of all the available evidence

Our findings should encourage public health officials and regulatory agencies to include the analysis of objective measures, in addition to self-reported questionnaires, as part of the evaluation of clinical trials. Though not available in the past, current digital wearable health technologies could offer simple platforms for obtaining physiological measures, adding invaluable objective, non-biased information. Thus, as part of the future process of vaccine approval, it is essential to combine objective and remote medical-grade monitors in clinical studies.

## Data Availability

Researchers interested in obtaining an aggregated version of the data sufficient to reproduce the results reported in this paper should contact the corresponding author. The study protocol is available at the appendix (pp 25-35). Statistical code will be available with publication.

## Authors’ Contributions

Conception and design: DY, ES, YG, Collection and assembly of data: MM, SO. Analysis and interpretation of the data: MM, SO, YG, ES, DY. Statistical expertise: DY, ES, MM, SO, MY. Drafting of the article: DY, ES, YG, MM, SO, MY, KC, NG. Critical revision of the article for important intellectual content: ES, DY, YG. Final approval of the article: All authors. Obtaining of funding: DY, ES.

## Supplementary Materials

### Appendix A: Study Protocol

#### -Study protocol-

(protocol approved by the Tel Aviv University Institutional Review Board: 0002522-1)

**Primary funding: the European Research Council (ERC) project #949850**

**Ethical Approval: Tel-Aviv University Institutional Review Board (0002522-1) December, 2020**

### Background

Vaccination is widely accepted as the most prominent measure in the fight against COVID-19, posing the greatest hope for ending this major global health pandemic and related economic crisis.^1^ Consequently, an unprecedented international effort by private and public institutions alike was directed at accelerating the traditionally lengthy vaccine-development process.^2–4^

On 2 December 2020, less than a year from the pandemic outbreak, the first vaccine, BNT162b2 mRNA (Pfizer-BioNTech), was granted an Emergency Use Authorization (EUA) by the UK Medicines and Healthcare products Regulatory Agency (MHRA).^5^ This initial authorization was followed by rapid authorizations for emergency use in several countries, with the US Food and Drug Administration (FDA) among the first to do so.^6^

Safety data from a randomized controlled trial suggests a favorable safety profile for the BNT162b2 vaccine.^7^ Specifically, the local and systemic self-reported reactions during the first seven days after vaccination were mainly mild to moderate, with a median onset of 0–2 days after vaccine administration and a median duration of 1–2 days. The most frequently reported reactions were fatigue, headache, muscle pain, chills, joint pain, and fever.^7^ The incidence of serious adverse events was low and was similar between vaccine- and placebo-treated participants. The safety of the new vaccine over a median of two months post-vaccination was similar to that of other viral vaccines. A considerable fraction of the participants did not report any reaction or adverse event. Likewise, several other vaccine candidates, including ChAdOx1 nCoV-19 (Oxford/AstraZeneca) and mRNA-1273 (Moderna), received EUAs following similar encouraging safety results by randomized controlled trials.^8–10^

Nevertheless, concerns regarding potential adverse effects from vaccines have recently led to the suspension of the ChAdOx1 nCoV-19 vaccination campaigns in Europe.^11^ They may have reinforced the public hesitancy towards COVID-19 vaccines. These concerns underscore the importance of extracting as much information as possible from clinical trials.

The World Health Organization (WHO) has defined vaccine hesitancy as one of the 10 major health threats worldwide.^12^ However, clinical trial guidelines for assessing the safety of vaccines, including the FDA criteria,^13^ are primarily rely on subjective, self-reported questionnaires. Despite the tremendous technological advances in recent years, objective, continuous assessment of physiological measures post-vaccination is rarely performed.

This short-term prospective clinical study aims to document symptoms among COVID-19 vaccinated subjects and cross it with physiological signs derived from continuous remote monitoring before, during, and after receiving the vaccine, looking at any potential hidden layers of the safety of the vaccine.

### Methods

#### Study Design

This prospective observational study will be conducted during the mass-vaccination campaign expected to be implemented in Israel in early 2021. The study design is outlined in Figure S1. Participants will be equipped with a chest-patch sensor and will be monitored for four days, starting one day before vaccination. In addition, participants will install a dedicated mobile application and will be requested to fill a daily questionnaire, starting one day before the inoculation, for 15 days. For each participant, the 24-hour period prior to vaccination will be served as the baseline period.

**Figure S1.**
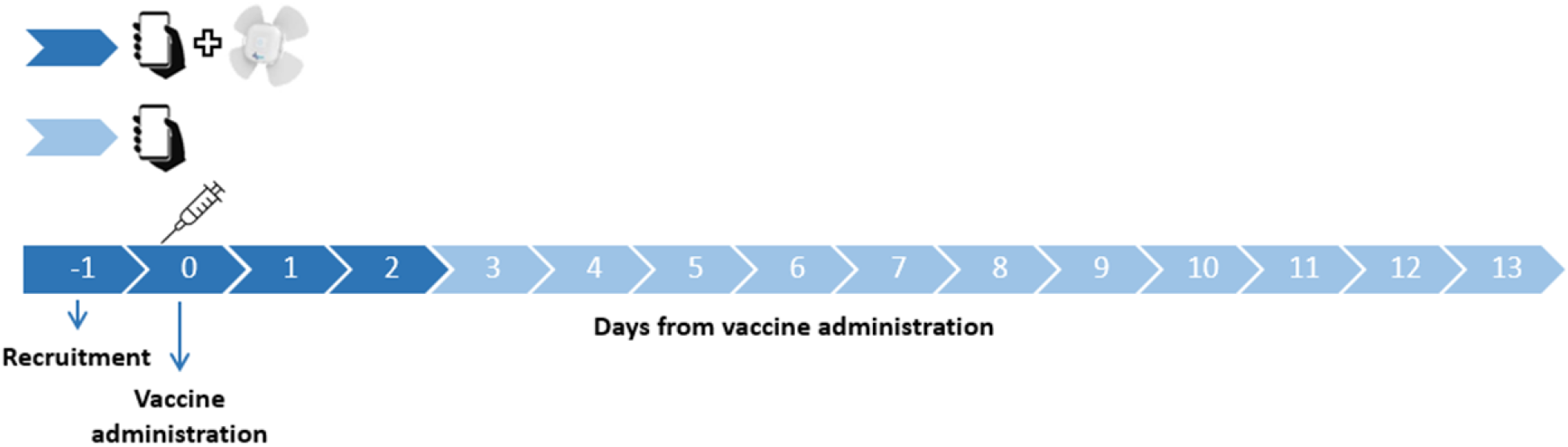
Study scheme.

#### Participants

Up to 200 participants will be recruited for the study, all before receiving the BNT162b2 mRNA COVID-19 vaccine. Inclusion criteria will include those aged > 18 years who were eligible to receive the vaccine. Individuals who are not eligible to give and sign a consent form of their free will or those previously diagnosed with COVID-19 will be excluded. To recruit participants and ensure they complete all the study’s requirements, we will hire a professional survey company. Potential participants will be recruited through advertisements in social media, online banners, and word-of-mouth. The survey company is responsible for guaranteeing the participants meet the study’s requirements, in particular, that they agree to wear the chest-patch sensor and fill in the daily questionnaires.

#### Study procedures

Several days before receiving the vaccine, a study investigator personally meets with each participant to explain the study procedure. Before participation in the study, all participants will be advised orally and in writing about the nature of the experiments and give written, informed consent. At this time, participants will be asked to complete enrollment questionnaire that includes demographic information and health status. In addition, participants will be asked to install two applications on their mobile phones: an application that passively collects data from the chest-patch sensor and the PerMed application, which allows participants to fill in the daily questionnaires. Participants will be given instructions regarding the self-reported symptoms questionnaires and how to operate the wireless chest-patch sensor, which they will wear for four consecutive days, starting 24 hours prior to receiving the vaccine.

#### Enrollment questionnaire

As mentioned, all the participants filled a one time enrollment questionnaire that includes demographic questions and questions about the participant’s health condition in general. The questionnaire we will use includes the following: age, gender, height, weight, household size and comorbidities. Other questions such as name, address, phone and email were included as well and were used by the survey company in order to contact the participants. The answers were inserted by the study investigator to the study’s secured dashboard.

#### Monitoring device

Starting 24 hours prior to receiving the vaccine, participants will wear a chest-patch sensor attached, continuously measuring 13 physiological parameters: heart rate, blood oxygen saturation, respiratory rate, systolic and diastolic blood pressure, pulse pressure, mean arterial pressure, heart rate variability, stroke volume, cardiac output, cardiac index, systemic vascular resistance, and body temperature. Participants will wear the monitoring sensor for three days following the vaccine.

The photoplethysmography (PPG)-based chest monitors (Biobeat Technologies Ltd) that will be purchased and used in this study has FDA clearance (clearance number K190792) and CE Mark approval (CE 2797). The sensor tracks vital signs derived from changes in the pulse contour, following baseline calibration using an approved non-invasive, cuff-based device, and is based on Pulse Wave Transit Time (PWTT) technology, combined with Pulse Wave Analysis (PWA). The sensor will continuously collect data at 10-minute intervals for the entire duration of the 96-hour experiment.

#### Daily questionnaires

All participants will complete the daily self-reported questionnaire in a dedicated application (the PerMed mobile application). The daily questionnaire we will use includes the following questions:

How is your mood today? • Awful (−2)• Bad (−1)• OK (0)• Good (1)• Excellent (2)

How would you describe the level of your stress during the last day?• Very Low (−2)• Low (−1)• Medium (0)• High (1)• Very high (2)

How would you define your last night sleep quality?• Awful (−2)• Bad (−1)• OK (0)• Good (1)• Excellent (2)

Try to remember how many minutes of sports activity you performed on the last day?

Have you experienced one or more of the following symptoms in the last 24 hours?• My general feeling is good, and I have no symptoms• Heat measured above 37.5• Cough• Sore throat• Runny nose• Headache• Shortness of breath• Muscle aches• Weakness / fatigue• Diarrhea• Nausea / vomiting• Chills• Confusion• Loss of sense of taste / smell• Another symptom

#### Data Storage

The data will be exported securely through the MD Clone secure data platform or a secured locked database only after receipt of consent forms from participants. Data from the mobile phone application of the prospective study population will be stored on a secure server within Tel Aviv University facilities. The server (bdl7.eng.tau.ac.il) runs a CentOS operating system and is located in Software Engineering Building at Tel Aviv University. This server is protected behind the university’s firewall and is not connected to external networks. In addition, a secure connection through an SSL protocol and a trusted certificate will be obtained for the transfer of information from the mobile phone application into the secured server.

Access will be restricted to investigators in the study. The information from the mobile application will be stored in a structured manner on the secured server without any explicitly identifying information (name, ID number, email). Each participant will be assigned a coded participant number that will be used to identify the subject in the database. The code with the identified information will be stored in an encrypted form on a separate secured server that only the research manager will have access to. Access to all servers is restricted with username and password.

All (non-digital) questionnaires and signed informed consent documents will be stored in a secured cabinet in Tel Aviv University, to which only the research manager and the principal investigators will have access.

No data collected as part of the study will be added to individuals’ medical charts.

All signed informed consent and printed questionnaires will be stored in a secured cabinet at Tel Aviv University with restricted access. Only the PIs will have access to this cabinet.

All individual clinical test results that are generated as part of this study will be shared with participants as explained above.

#### Data processing

We will perform several preprocessing steps. Concerning the daily questionnaires, in cases where participants will fill in the daily questionnaire more than once on a given day, only the last entry for that day will be considered, as it is reasoned that the last one likely best represented the entire day. Self-reported symptoms that are entered as the free text will be manually categorized. Concerning the chest-patch indicators, data will first be aggregated per hour (by taking the mean value). Then, to impute missing values, we will perform a linear interpolation.

#### Data Analysis

We will examine the changes in each chest-patch indicator over the 72 hours post-vaccination compared with the 24-hour baseline period prior to vaccination. To do so, we will perform the following steps. For each indicator, for each hour *h* of the 72 hours post-vaccination, we will calculate for each individual the mean value in the five-hour sliding window: [*h* − 4, *h* − 3, *h* − 2, *h* − 1, *h*]. Then, we will calculate the relative change in the percentage of this value compared to the corresponding five-hour window in the baseline period. Finally, we will calculate the mean value for hour *h* over all 160 participants, as well as the 90% confidence interval, corresponding to a significance level of 0.05 in a one-sided t-test.

We will also examine the changes in the daily questionnaire. For each of the three days after the vaccination, we will calculate the percentage of participants who reported new local or systemic reactions compared to their baseline period. For each reaction, a 90% confidence interval will be calculated assuming a beta distribution, with parameter *α* corresponding to the number of participants reporting that reaction plus one (i.e., “successes”), and parameter *β* corresponding to the number of participants who did not report that reaction plus one (i.e., “failures”). Similarly, we will calculate the changes in the well-being indicators reported post-vaccination compared to those reported in the baseline period. Specifically, we will calculate the difference between the value in each day and the corresponding value in the baseline period for each indicator. Then, we will calculate the mean difference value over all participants and a 90% confidence interval using a t distribution.

Finally, we will examine the difference between symptomatic and asymptomatic participants with regard to changes in the chest-patch indicators, stratified by the number of days post-vaccination (1-3) and part of the day (day or night). For a given day post-vaccination, symptomatic participants will be defined as those who reported at least one reaction on that day that they did not report in the baseline period. Asymptomatic individuals will be defined as those who reported no reactions on that day. We will define nighttime and daytime based on the observed movement patterns of the participants throughout the study. For each participant, we will calculate the mean indicator value for each day and part of the day post-vaccination. Then, we will calculate the relative change in percentages of these values compared to their corresponding values in the baseline period. Next, we will calculate the mean values of symptomatic participants and asymptomatic participants and their corresponding 90% confidence intervals using a t distribution. Finally, unequal variances t-tests will be used to evaluate the differences between symptomatic and asymptomatic participants.

#### Potential Risks & Risk management

No potential risks arising from the sensors are expected, as the device is already commercialized with no known adverse reactions. Following the instructions for use, people with known allergies to metals (e.g., zinc, tin) will not be included in the study.

#### Benefits

The sensor used in this study will provide continuous blood pressure data for each participant. After completion of the study, all participants will receive a detailed report containing information regarding their fluctuations in blood pressure during the baseline monitoring period (i.e., 24 hours prior to receiving the vaccine), with a recommendation to share it with their general practitioner.

#### Privacy/Confidentiality

Results from this study will be handled as grouped data. Individual results will remain confidential and will not be published or shared with any third party. Signed and dated informed consent forms, as well as data recording sheets (e.g., case report forms) will be stored in locked cabinets during the study and following its completion. A file containing the personal details of the participants will be coded to help preserve confidentiality and separated from all other data collected throughout the study. This file will be kept by the principal investigator. Data will be stored on computers in password-protected files.

The data obtained from the sensor used in this study will be anonymous and coded. The sensor does not include a GPS, as required from any medical-grade monitoring device. The data collected by the PerMed application arrive to PerMed back-end servers and are stored securely.

### Appendix B: Additional Results

#### Additional chest-patch indicators

Within the first 48 hours post-vaccination, we identified significant changes in nearly all chest-patch physiological measures compared to their baseline levels (Figure 1 in the main text and Figure S2). Following the initial 48 hours, these changes faded, with measurements returning to their baseline levels.

**Figure S2.**
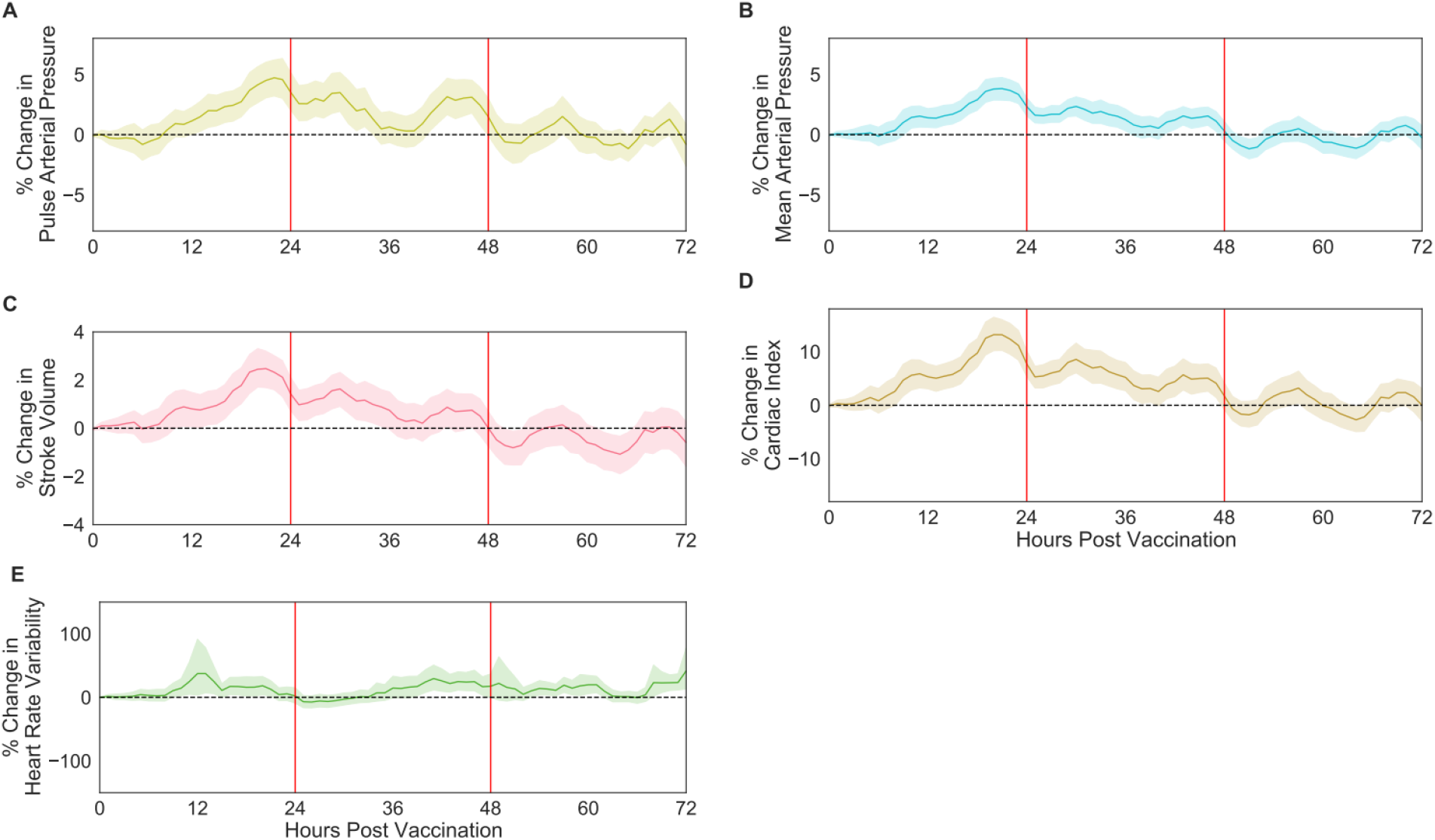
Percentage of change in respiratory, cardiovascular, and physiological chest-patch indicators recorded by the chest-patch sensor, compared to their baseline levels: (A) pulse arterial pressure, (B) mean arterial pressure, (C) stroke volume, (D) cardiac index, (E) heart rate variability. Mean values are depicted as solid lines, 90% confidence intervals are presented as shaded regions, and horizontal dashed lines represent no change compared to the baseline levels.

#### Additional days for self-reported local and systemic reactions

We observe a sharp decline in reported local and systemic reactions following three days post-vaccination and nearly a complete halt within 14 days post-vaccination (Figure 2 in the main text and Figure S3). Fatigue and headache were the most frequent reactions reported and lasted longer than the other reported reactions.

**Figure S3.**
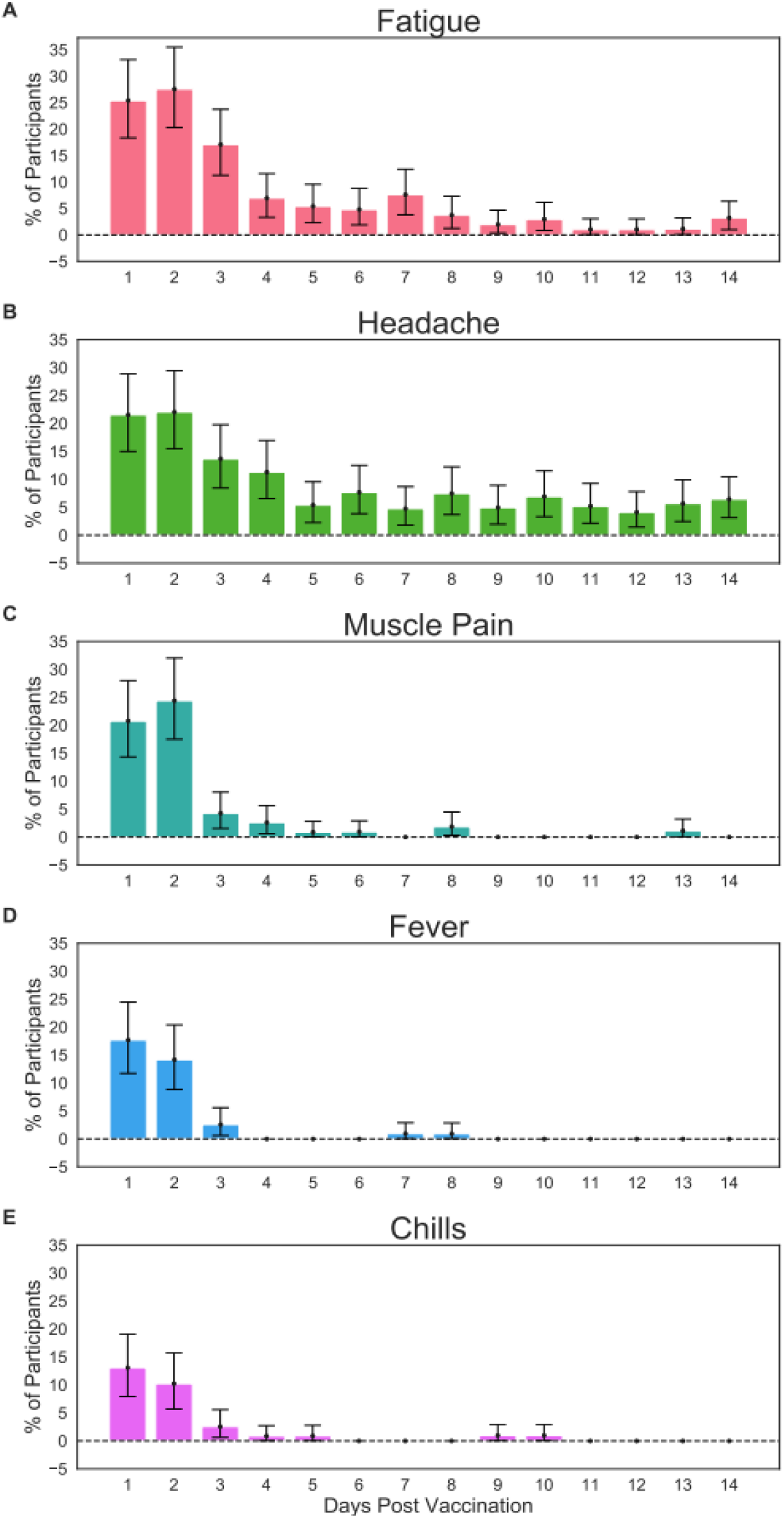
Most frequent local and systemic reactions reported by participants through the mobile application. (A) fatigue, (B) headache, (C) muscle pain, (D) fever, and (E) chills. Error bars represent 90% confidence intervals. Horizontal dashed lines represent no change compared to the baseline levels.

#### Symptomatic vs. asymptomatic participants – additional chest-patch indicators

During the daytime of the first two days post-vaccination, nearly all chest-patch indicators significantly changed not only for symptomatic participants but also for asymptomatic participants (Figure 3 in the main text and Figure S4). Moreover, the changes observed for symptomatic participants were found to be significantly higher than those of asymptomatic individuals in most chest-patch indicators during the first day and the first night post-vaccination.

**Figure S4.**
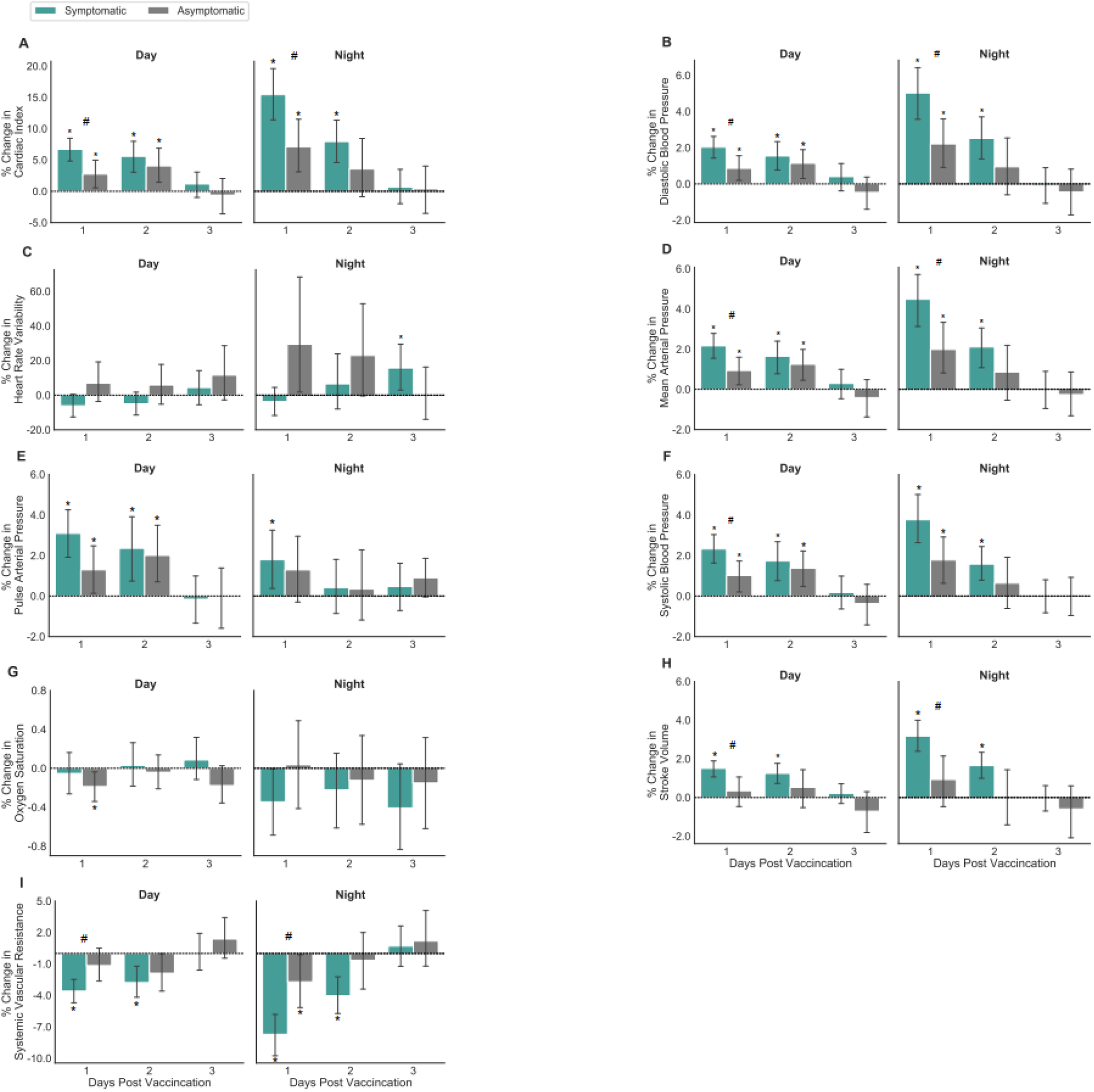
Percentage of change in chest-patch indicators for participants who reported at least one local or systemic reaction (symptomatic) and those who reported no reaction (asymptomatic), during days and nights: (A) cardiac index, (B) diastolic blood pressure, (C) heart rate variability, (D) mean arterial pressure, (E) pulse arterial pressure, (F) systolic blood pressure, (G) oxygen saturation, (H) stroke volume, and (I) systemic vascular resistance. Error bars represent 90% confidence intervals. Horizontal dashed lines represent no change compared to the baseline levels. Significant differences from baseline at a 0.05 level are marked with *. Significant differences between symptomatic and asymptomatic participants at a 0.05 level are marked with #.

#### Gender groups differences in chest-patch indicators

During the daytime and nighttime of the first two days post-vaccination, all four chest-patch indicators significantly changed for both males and females (Figure S5). There was no significant change between the two genders.

**Figure S5.**
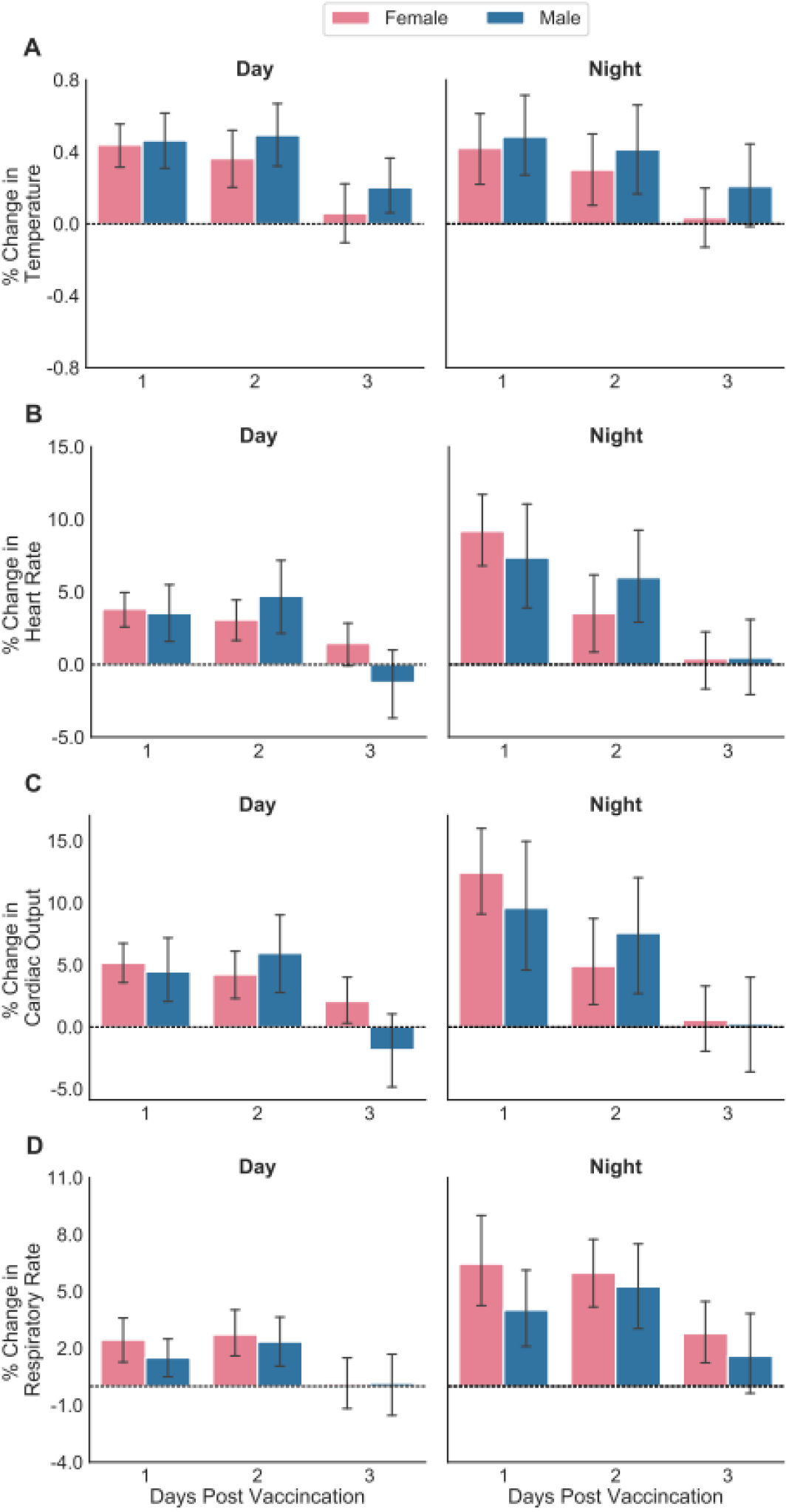
Percentage of change in chest-patch indicators for participants divided by gender during the daytime and the nighttime: (A) body temperature, (B) heart rate, (C) cardiac output, and (D) respiratory rate. Error bars represent 90% confidence intervals. Horizontal dashed lines represent no change compared to the baseline levels.

#### Age groups differences in chest-patch indicators

During the daytime and nighttime of the first two days post-vaccination, all four chest-patch indicators significantly changed for the younger age group but not necessary for the older group (Figure S6).

**Figure S6.**
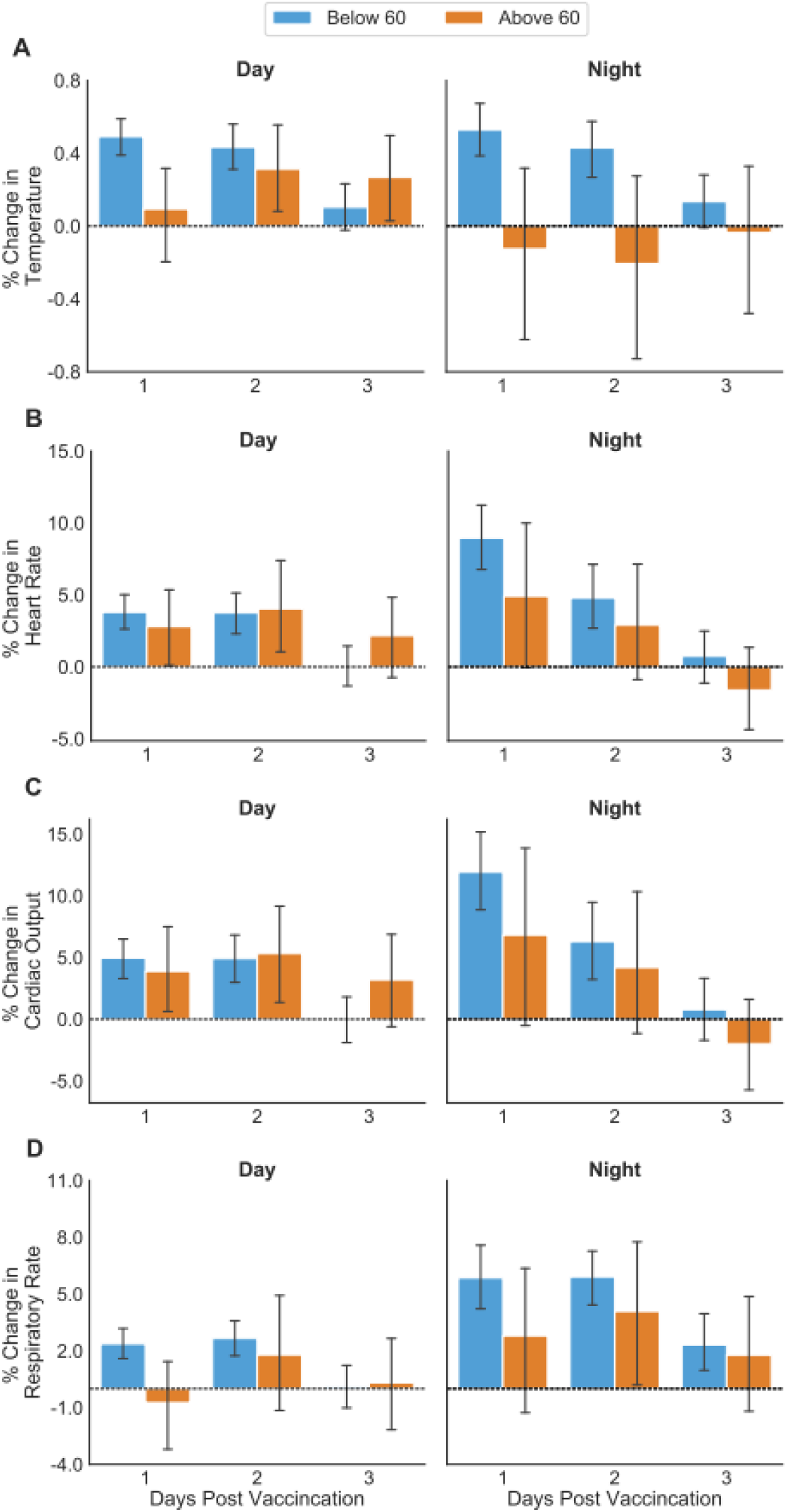
Percentage of change in chest-patch indicators for participants divided by age group during the daytime and the nighttime: (A) body temperature, (B) heart rate, (C) cardiac output, and (D) respiratory rate. Error bars represent 90% confidence intervals. Horizontal dashed lines represent no change compared to the baseline levels.

#### Chest-patch indicators for first vaccine dose

In contrast to the second vaccine dose, examining the participants who were monitored when receiving their first vaccine dose revealed no difference in vitals during the first 48 hours post-inoculation.

**Figure S7.**
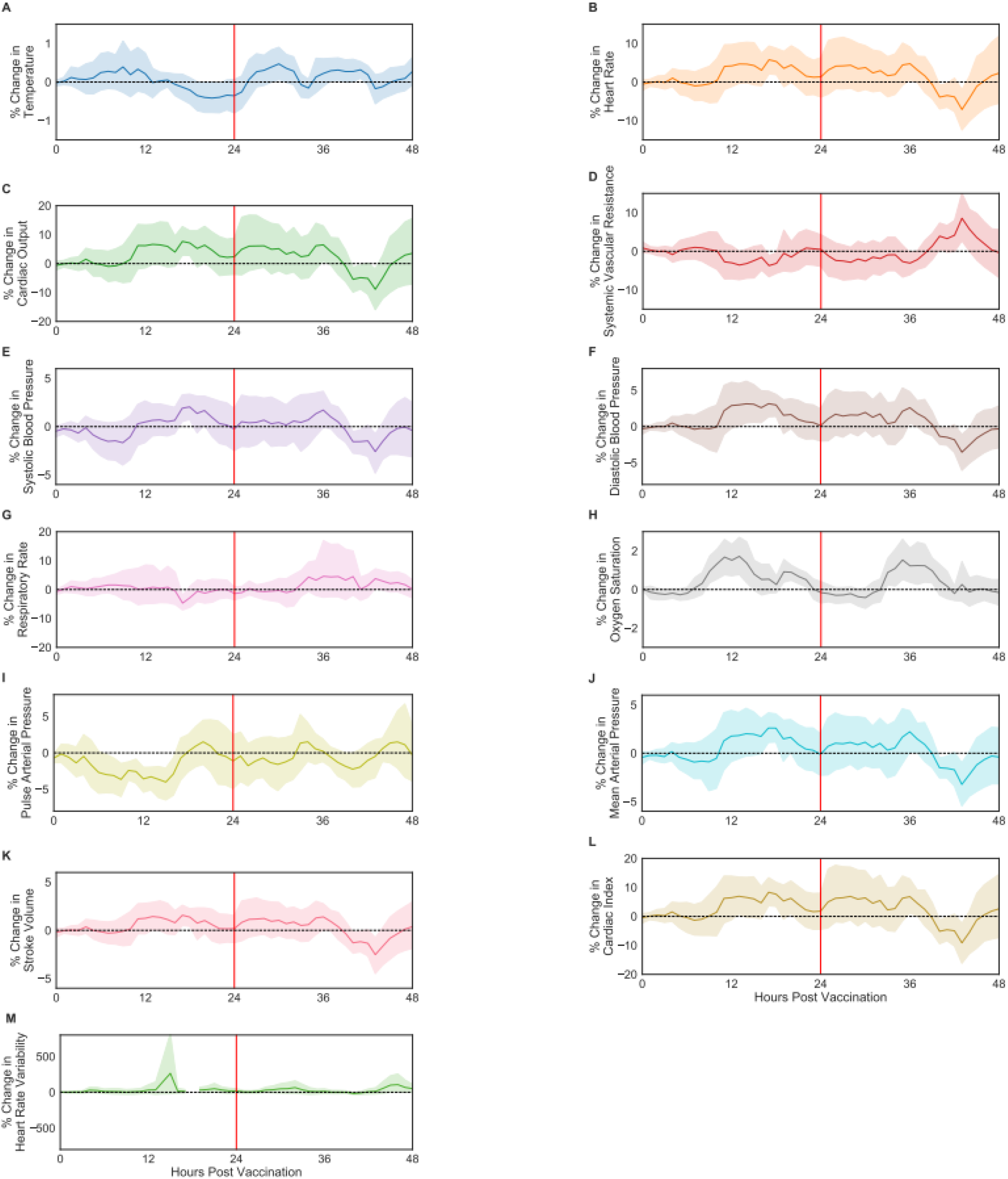
Percentage of change in respiratory, cardiovascular, and physiological indicators recorded by the chest-patch sensor compared to their baseline levels: (A) body temperature, (B) heart rate, (C) cardiac output, (D) systemic vascular resistance, (E) systolic blood pressure, (F) diastolic blood pressure, (G) respiratory rate, (H) oxygen saturation, (I) pulse arterial pressure, (J) mean arterial pressure, (K) stroke volume, (L) cardiac index, and (M) heart rate variability. Mean values are depicted as solid lines, 90% confidence intervals are presented as shaded regions, and horizontal dashed lines represent no change compared to the baseline levels.

## References

1 Draft landscape and tracker of COVID-19 candidate vaccines. https://www.who.int/publications/m/item/draft-landscape-of-covid-19-candidate-vaccines (accessed March 31, 2021).

2 Munitz A, Yechezkel M, Dickstein Y, Yamin D, Gerlic M. BNT162b2 vaccination effectively prevents the rapid rise of SARS-CoV-2 variant B.1.1.7 in high-risk populations in Israel. Cell Reports Med 2021; 0: 100264.

3 Sharma O, Sultan AA, Ding H, Triggle CR. A Review of the Progress and Challenges of Developing a Vaccine for COVID-19. Front Immunol 2020; 11: 585354.

4 Parker EPK, Shrotri M, Kampmann B. Keeping track of the SARS-CoV-2 vaccine pipeline. Nat. Rev. Immunol. 2020; 20: 650.

5 Krammer F. SARS-CoV-2 vaccines in development. Nature. 2020; 586: 516–27.

6 MHRA guidance on coronavirus (COVID-19) - GOV.UK. https://www.gov.uk/government/collections/mhra-guidance-on-coronavirus-covid-19 (accessed March 31, 2021).

7 FDA Takes Key Action in Fight Against COVID-19 By Issuing Emergency Use Authorization for First COVID-19 Vaccine | FDA. https://www.fda.gov/news-events/press-announcements/fda-takes-key-action-fight-against-covid-19-issuing-emergency-use-authorization-first-covid-19 (accessed March 31, 2021).

8 Polack FP, Thomas SJ, Kitchin N, et al. Safety and Efficacy of the BNT162b2 mRNA Covid-19 Vaccine. N Engl J Med 2020; 383: 2603–15.

9 Dagan N, Barda N, Kepten E, et al. BNT162b2 mRNA Covid-19 Vaccine in a Nationwide Mass Vaccination Setting. N Engl J Med 2021; : NEJMoa2101765.

10 Background document on mRNA vaccine BNT162b2 (Pfizer-BioNTech) against COVID-19. https://www.who.int/publications/i/item/background-document-on-mrna-vaccine-bnt162b2-(pfizer-biontech)-against-covid-19 (accessed March 31, 2021).

11 Baden LR, El Sahly HM, Essink B, et al. Efficacy and Safety of the mRNA-1273 SARS-CoV-2 Vaccine. N Engl J Med 2021; 384: 403–16.

12 Voysey M, Clemens SAC, Madhi SA, et al. Safety and efficacy of the ChAdOx1 nCoV-19 vaccine (AZD1222) against SARS-CoV-2: an interim analysis of four randomised controlled trials in Brazil, South Africa, and the UK. Lancet 2021; 397: 99–111.

13 Mahase E. Covid-19: WHO says rollout of AstraZeneca vaccine should continue, as Europe divides over safety. BMJ 2021; 372: 728.

14 Development and Licensure of Vaccines to Prevent COVID-19 | FDA. https://www.fda.gov/regulatory-information/search-fda-guidance-documents/development-and-licensure-vaccines-prevent-covid-19 (accessed March 31, 2021).

15 Nachman D, Gepner Y, Goldstein N, et al. Comparing blood pressure measurements between a photoplethysmography-based and a standard cuff-based manometry device. Sci Rep 2020; 10: 1–9.

16 Halberthal M, Nachman D, Eisenkraft A, Jaffe E. Hospital and home remote patient monitoring during the COVID-19 outbreak: A novel concept implemented. Am J Disaster Med 2020; 15: 149– 51.

17 Oved S, Mofaz M, Lan A, et al. Differential effects of COVID-19 lockdowns on well-being: interaction between age, gender and chronotype. 2021 DOI:10.21203/RS.3.RS-137929/V1.

18 Non-FDA. Vaccines and Related Biological Products Advisory Committee December 10, 2020 Meeting Briefing Document-Sponsor. 2020.

## References

2 Sharma O, Sultan AA, Ding H, Triggle CR. A Review of the Progress and Challenges of Developing a Vaccine for COVID-19. Front Immunol 2020; 11: 585354.

3 Parker EPK, Shrotri M, Kampmann B. Keeping track of the SARS-CoV-2 vaccine pipeline. Nat. Rev. Immunol. 2020; 20: 650.

4 Krammer F. SARS-CoV-2 vaccines in development. Nature. 2020; 586: 516–27.

5 MHRA guidance on coronavirus (COVID-19) - GOV.UK. https://www.gov.uk/government/collections/mhra-guidance-on-coronavirus-covid-19 (accessed March 31, 2021).

6 FDA Takes Key Action in Fight Against COVID-19 By Issuing Emergency Use Authorization for First COVID-19 Vaccine | FDA. https://www.fda.gov/news-events/press-announcements/fda-takes-key-action-fight-against-covid-19-issuing-emergency-use-authorization-first-covid-19 (accessed March 31, 2021).

7 Polack FP, Thomas SJ, Kitchin N, et al. Safety and Efficacy of the BNT162b2 mRNA Covid-19 Vaccine. N Engl J Med 2020; 383: 2603–15.

8 Background document on mRNA vaccine BNT162b2 (Pfizer-BioNTech) against COVID-19. https://www.who.int/publications/i/item/background-document-on-mrna-vaccine-bnt162b2-(pfizer-biontech)-against-covid-19 (accessed March 31, 2021).

9 Baden LR, El Sahly HM, Essink B, et al. Efficacy and Safety of the mRNA-1273 SARS-CoV-2 Vaccine. N Engl J Med 2021; 384: 403–16.

10 Voysey M, Clemens SAC, Madhi SA, et al. Safety and efficacy of the ChAdOx1 nCoV-19 vaccine (AZD1222) against SARS-CoV-2: an interim analysis of four randomised controlled trials in Brazil, South Africa, and the UK. Lancet 2021; 397: 99–111.

11 Mahase E. Covid-19: WHO says rollout of AstraZeneca vaccine should continue, as Europe divides over safety. BMJ 2021; 372: 728.

12 Ten threats to global health in 2019. https://www.who.int/news-room/spotlight/ten-threats-to-global-health-in-2019 (accessed May 2, 2021).

13 Development and Licensure of Vaccines to Prevent COVID-19 | FDA. https://www.fda.gov/regulatory-information/search-fda-guidance-documents/development-and-licensure-vaccines-prevent-covid-19 (accessed March 31, 2021).

